# Cell type specific enrichment of substance use and substance use disorder heritability

**DOI:** 10.64898/2026.10.02.26364595

**Authors:** Nithya Sarabudla, Pamela N Romero Villela, Ronald P. Hart, Yang E. Li, Zhiping P. Pang, Arpana Agrawal, Emma C Johnson

**Affiliations:** Department of Psychiatry, Washington University School of Medicine, Saint Louis, MO, USA; Department of Cell Biology and Neuroscience, Rutgers University, Piscataway, NJ, USA; Department of Neurosurgery, Washington University School of Medicine, Saint Louis, MO, USA; Department of Genetics, Washington University School of Medicine, Saint Louis, MO, USA; Department of Neuroscience and Cell Biology, Rutgers Robert Wood Johnson Medical School, New Brunswick, NJ, USA

## Abstract

Genome-wide association studies (GWAS) have identified numerous loci associated with substance use disorders (SUDs). To connect common genetic variation to cell type- and region-specific gene expression and chromatin accessibility, we examined the enrichment of genetic liability for several SUDs (alcohol, cannabis, tobacco, and opioid use disorders) and substance consumption phenotypes (drinks per week, cannabis ever-use, and cigarettes per day) in available single-nucleus RNAseq data spanning 10 brain regions and snATACseq data spanning 42 brain regions. Significant cell-type enrichment after multiple testing corrections was observed only for drinks per week and tobacco use disorder in the snRNAseq data and for cannabis use disorder and problematic alcohol use in snATACseq data. Across both transcriptomic and chromatin accessibility annotations, enrichment patterns were concentrated in excitatory neuronal populations, particularly upper layer intratelencephalic neurons, amygdala excitatory neurons, and regionally resolved striatal medium spiny neuron subtypes. Drinks per week, problematic alcohol use, and cannabis use disorder showed enrichment values that were significantly correlated within cell-type across snRNAseq and snATACseq, suggesting convergent biological signals across independent single-nucleus modalities. These findings implicate chromatin-mediated expression differences in specific excitatory neuronal populations as potential mediators of SUD genetic liability and highlight the value of integrating transcriptomic and chromatin accessibility data for characterising the neurobiology of addiction.

## Introduction

Substance use behaviors and substance use disorders (SUDs) are heritable (twin-h^2^ = 30 - 70%, SNP-h², 5 - 15%), multifactorial phenotypes, with both environmental and genetic factors shaping individual differences^1^. Genome-wide association studies (GWAS) of SUDs have identified hundreds of common genetic variants correlated with risk for substance use and SUDs, implicating some substance-specific genes (e.g., alcohol metabolizing genes, such as *ADH1B*) as well as genes that seem to be shared across SUDs (e.g., *DRD2* and *PDE4B)*^2^. Interestingly, while substance use and SUD genetic signals have been found to be highly correlated in aggregate (e.g., SNP-r_g_(drinks per week, problematic alcohol use) = 0.77; SNP-r_g_(cannabis ever-use, cannabis use disorder) = 0.50), there is also support for some distinctions between genetic factors associated with lifetime substance use and those contributing to progression to disordered use. For instance, while drinks per week and cannabis ever-use genetically correlate with higher educational attainment and socioeconomic status, the genetics of alcohol and cannabis use disorders document opposing effects^3,4^, suggesting partially distinct etiologies.

Notably, for both substance use and SUDs, as for other psychiatric traits, there is broad consensus for enrichment of common loci in regulatory regions and for genome-wide significant variants mapping to brain cis-eQTLs rather than directly modifying transcription^5,6^. The association of GWAS signals with inferred variation in gene expression could be due to various mechanisms that regulate expression, some of which may be tissue- and cell-type specific. For instance, a large GWAS examining cross-SUD genetic liability found chromatin-folding as a key regulatory mechanism underlying the dopaminergic genetic signals that emerged in the study^2^. Yet, few studies have examined co-enrichment of GWAS signals across multiple regulatory modalities in a cell-type-specific manner. With the advent of single-nucleus multiome arrays (i.e., characterization of both chromatin accessibility peaks and gene expression in the same nuclei), chromatin accessibility has emerged as a potential contributor to cell-type specific gene expression differences, with one study finding that widespread gene expression differences in diverse cell types coincided with corresponding chromatin accessibility differences in the same genes in the caudate nucleus of individuals with alcohol use disorder^7^. However, such assays of post-mortem data from individuals with a history of chronic substance exposure can conflate the epigenetic effects of exposure and it correlates with predispositional regulatory variation.

Consistent with this, few post-mortem gene expression studies that compare samples from decedents with and without a history of SUD have found enrichment of GWAS signals for the corresponding SUD. This suggests that alternative methods for aligning GWAS signals with gene expression and chromatin accessibility data from diverse brain samples without a specific history of substance exposure may provide biological insights into predispositional regulatory mechanisms that confer liability to substance use and SUDs.

In the current study, we leveraged recently published single-nucleus gene expression (i.e., RNA-seq) and chromatin accessibility (i.e., ATAC-seq) data from up to 42 brain regions derived from human brain samples to assess cell type-specific enrichment of SUD and substance use behaviour heritability. We analyzed multiple SUDs (alcohol, cannabis, tobacco, and opioid use disorders) and substance use phenotypes (drinks per week, cannabis ever-use, and cigarettes per day). We also characterised the degree to which snRNAseq and snATACseq enrichments were correlated within the same region and cell types to determine whether our findings might be consistent with chromatin accessibility as a potential mechanism linking GWAS loci with variability in gene expression.

## Materials and methods

### Genome-wide association studies of substance use disorders

Seven publicly available summary statistics from genome-wide association studies (GWAS) of substance use and SUD phenotypes were obtained. We restricted our analyses to GWAS of European ancestry subsamples, as these GWAS had the largest sample sizes, and the LDSC enrichment analyses must be performed within ancestry due to differences in linkage disequilibrium (LD) across populations. These GWAS included cannabis use disorder (CanUD; Levey et al., 2023^8^), cannabis ever-use (Bright et al., 2026^9^), opioid use disorder (OUD; Deak et al., 2022^10^), problematic alcohol use (PAU; Zhou et al., 2023^11^), drinks per week (Saunders et al., 2022^12^), cigarettes per day (CPD; Saunders et al., 2022^12^), and tobacco use disorder (TUD; Toikumo et al., 2024^1^) **(Table S1)**. Additionally, Alzheimer’s disease GWAS summary statistics (Wightman et al. 2021^13^) were included as a negative control phenotype, as it is a neuropsychiatric disorder with a predominantly distinct genetic architecture (with null to small genetic correlations with SUDs: r_g_s = −0.03 to 0.18; **Table S2**), allowing us to assess the specificity of the SUD enrichment signals. For case-control phenotypes, we calculated effective sample size as 4 / (1/N cases + 1/N controls). Prior to enrichment analysis, summary statistics for all seven phenotypes were processed using the munge_sumstats.py script from the LDSC software package, using the 1000 Genomes European ancestry reference panel and filtering to HapMap3 SNPs.

### Single-cell RNAseq and ATACseq data

Single-nucleus RNA sequencing (snRNAseq) data were obtained from Siletti et al. (2023)^14^, a comprehensive transcriptomic atlas of the adult human brain encompassing 31 broad cell-type supercluster annotations derived from ∼3 million nuclei across 105 dissections spanning 10 brain regions from 4 postmortem donors, profiled using 10x Genomics Chromium v3 chemistry.

Single-nucleus ATAC sequencing (snATACseq) data were obtained from Li et al. (2023)^15^, a chromatin accessibility atlas of the adult human brain comprising 109 cell types characterised from ∼1.1 million nuclei across 42 brain regions from 3 neurotypical postmortem donors, profiled using snATAC-seq. To enable meaningful comparison with the 31 snRNAseq supercluster annotations in downstream analyses, the 109 snATACseq cell types were consolidated into 49 broader cell classes based on shared cell-type identity and regional origin (**Table S3**).

### Testing for enrichment using linkage disequilibrium score regression

Cell-type-specific heritability enrichment was estimated using the --h2-cts option of the LDSC software^16^. For each phenotype, LDSC was run conditioned on the baseline v1.2 annotation model, using 1000 Genomes Phase 3 European ancestry LD scores as the reference panel and HapMap3 SNPs (excluding the major histocompatibility complex (MHC) region) as regression weights. Enrichment was tested separately for the snRNAseq and snATACseq annotations across all seven substance use phenotypes and our negative control phenotype. Statistical significance was assessed using a Benjamini-Hochberg false discovery rate correction (FDR < 0.05) applied within each substance use phenotype, as well as a more stringent Bonferroni correction (*p* < 0.05 / number of cell types tested), both applied separately for each annotation system (Bonferroni *p* < 1.61 × 10 ³ for 31 snRNAseq superclusters; *p* < 1.02 × 10 ³ for 49 snATACseq cell types).

### *Estimating correlations between snRNAseq and snATACseq enrichment and between* substance use and use disorders

To assess the concordance between the snRNAseq and snATACseq enrichment results, we computed Pearson correlations between enrichment Z-scores across matched cell types for each SUD phenotype. Cell-type matching was performed using curated mapping between snATACseq consolidated cell classes and snRNAseq superclusters, based on shared cell-type identity and regional origin (**Table S4**). For snATACseq cell types mapping to multiple snRNAseq supercluster categories, Z-scores were averaged within each consolidated category prior to matching. snATACseq cell types lacking a clear equivalent in the snRNAseq data were excluded from analyses. Cell-type matching resulted in 44 paired observations per phenotype. Pearson correlations and associated *p*-values were computed using the cor.test() function in R (version 4.2.3).

To determine whether within-phenotype cross-modality correlations were greater than expected based on cross-phenotype concordance, we additionally calculated Pearson correlations for all pairwise combinations of the seven phenotypes across the two annotation modalities. The off-diagonal correlations, representing mismatched phenotype pairs, were used to characterize the background distribution. These correlations were Fisher z-transformed, and a normal distribution was fitted to the transformed values. Normality of the Fisher *z*-transformed correlations was assessed using the Shapiro-Wilk test. The 95th and 99th percentiles of the fitted distribution were calculated and back-transformed to the Pearson correlation scale to provide reference thresholds for evaluating the seven matched within-phenotype correlations.

To evaluate the similarity between genetic enrichment profiles of SUDs and corresponding substance use phenotypes, we also computed Pearson correlations between enrichment Z-scores for the same cell types across each disorder-use phenotype pair within each annotation modality. Correlations were calculated separately for TUD and cigarettes per day, PAU and drinks per week, and CanUD and cannabis ever-use in both snRNA-seq and snATAC-seq annotations. OUD was excluded because no corresponding substance use phenotype was available.

## Results

### Cell-type enrichment of SUD heritability in snRNAseq annotations

The cell-type-specific enrichment analysis of seven substance use and SUD GWAS in the snRNAseq supercluster annotations revealed a consistent directional pattern: enrichment coefficients were predominantly positive in neuronal populations and negative in glial and non-neuronal cell types, including Oligodendrocyte, Astrocyte, Microglia, Vascular, and Ependymal superclusters, which showed negative coefficients across the majority of phenotypes (**Figure 1**).

**Figure 1:**
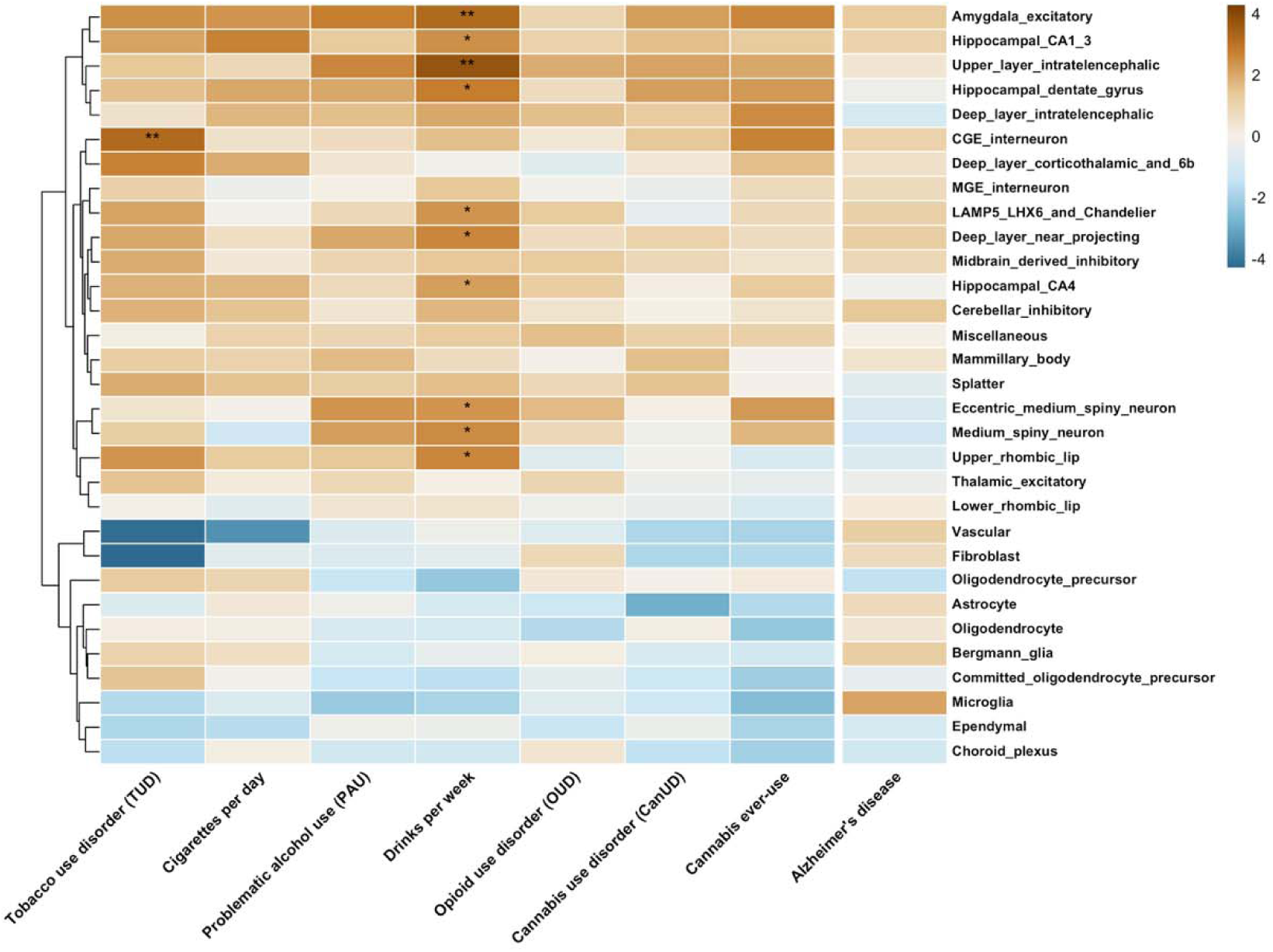
LDSC enrichment Z-scores across 31 snRNAseq cell-type superclusters for seven substance use phenotypes identified brain region–cell type-specific enrichment patterns for Drinks per Week and Tobacco Use Disorder. Brown indicates positive enrichment; blue indicates negative enrichment. Rows are hierarchically clustered. * indicates FDR < 0.05; ** indicates associations significant after both FDR and Bonferroni correction (Bonferroni *p* < 1.61 × 10⁻³). Among the seven phenotypes, drinks per week and tobacco use disorder exhibited the strongest cell-type-specific enrichment but with distinct patterns: drinks per week showed broad enrichment across excitatory neuronal populations, whereas tobacco use disorder was uniquely enriched in an inhibitory interneuron population. In contrast, Alzheimer’s disease, included as a negative control, showed a distinct pattern, with positive enrichment in the microglial annotation.

After FDR correction (Benjamini-Hochberg, FDR < 0.05) was applied within each phenotype, significant enrichment was observed for two phenotypes. Drinks per week showed the most widespread FDR-significant enrichment, with 10 cell types surviving correction, most prominently upper layer intratelencephalic (Z = 3.73, p = 9.68 × 10, FDR-adjusted p = 0.003) and amygdala excitatory (Z = 3.22, p = 6.38 × 10, FDR-adjusted p = 0.010) neurons. TUD yielded a single FDR-significant association with CGE interneuron (Z = 3.19, p = 7.21 × 10, FDR-adjusted p = 0.022), a GABAergic inhibitory interneuron population, which contrasts with the predominantly excitatory neuronal enrichment observed across other phenotypes. Three of these 11 FDR-significant cell-type enrichment associations also survived the more stringent Bonferroni correction (p < 1.61 × 10 ³): upper layer intratelencephalic and amygdala excitatory neurons for drinks per week, and CGE interneuron for TUD (**Figure 2**).

**Figure 2.**
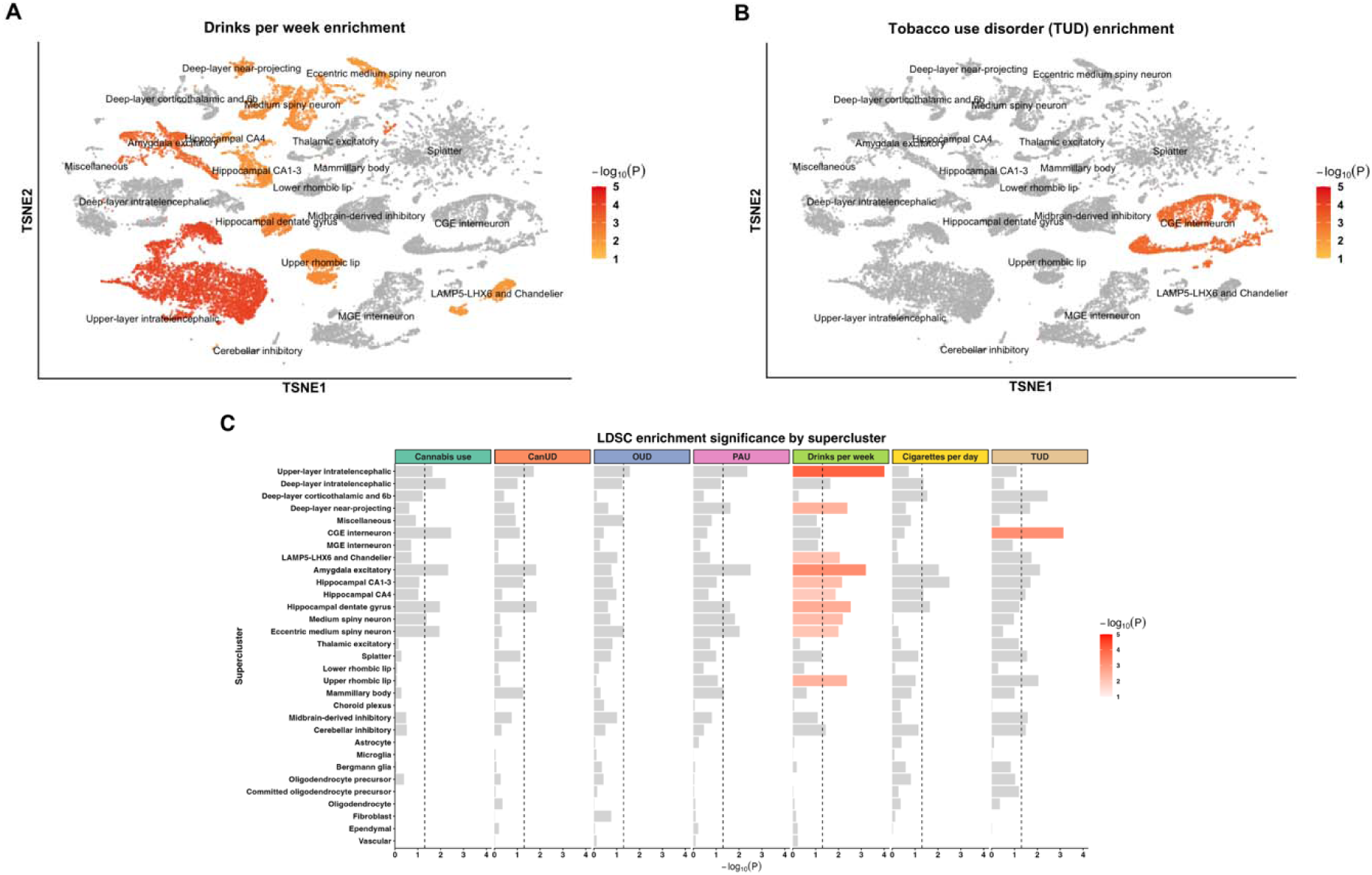
Amygdala excitatory and upper-layer intratelencephalic neurons showed the strongest enrichment for drinks per week (A), whereas CGE interneurons-a GABAergic population, were the only significantly enriched cell type for tobacco use disorder (TUD) (B), highlighting distinct cell-type-specific enrichment patterns between the two phenotypes with FDR-significant enrichment. A) t-Distributed Stochastic Neighbour Embedding (tSNE) plot of Siletti et al. snRNA-seq superclusters colored by −log10(*P*) enrichment values for drinks per week. Grey indicates non-significantly enriched superclusters (FDR > 0.05). B) tSNE plot of Siletti et al. snRNA-seq superclusters colored by −log10(*P*) enrichment values for TUD. Grey indicates non-significantly enriched superclusters (FDR > 0.05). C) Bar plot showing LDSC enrichment significance across 31 snRNA-seq superclusters for seven SUD phenotypes. Bars are colored by −log10(*P*) for superclusters surviving FDR correction (FDR < 0.05) and grey otherwise. The dashed vertical line indicates the nominal significance threshold (*p* < 0.05).

Looking across all enrichments with p < 0.05 (uncorrected for multiple testing), a broader and largely consistent pattern emerged. The amygdala excitatory annotation was the most consistently enriched across phenotypes, reaching nominal significance for six of seven (drinks per week, PAU, CanUD, cannabis use, cigarettes per day, and TUD). The upper-layer intratelencephalic annotation was enriched for five phenotypes (drinks per week, PAU, CanUD, cannabis use, and OUD), and the hippocampal dentate gyrus annotation for five as well (drinks per week, PAU, cigarettes per day, CanUD, and cannabis use). Striatal populations showed similar recurrence: the eccentric medium spiny neuron annotation was enriched for four phenotypes (drinks per week, PAU, cannabis use, and OUD), and the medium spiny neuron annotation for three (drinks per week, PAU, and cannabis use), consistent with the established role of striatal circuitry in reward processing and addiction. Overall, neuronal annotations showed recurrent positive enrichment across multiple phenotypes, suggesting shared patterns of cell-type-specific heritability enrichment across substance use and SUD phenotypes. As a negative control, the same annotations tested against Alzheimer’s disease heritability showed a markedly distinct profile, with enrichment concentrated in the microglial annotation rather than excitatory neuronal annotations (**Figure 1**), supporting the specificity of the SUD signal reported here. Although neuronal pathology is prominent in postmortem Alzheimer’s disease tissue, the microglial enrichment observed here is consistent with existing evidence implicating microglia in Alzheimer’s genetic susceptibility^17^.

### Cell-type enrichment of SUD heritability in snATACseq annotations

The cell-type-specific enrichment analysis in the snATACseq annotations, comprising 109 cell types consolidated into 49 broader cell classes, revealed a broadly consistent directional pattern with the snRNAseq results: positive enrichment in neuronal cell types and negative enrichment in non-neuronal cell types (**Figure 3**). After FDR correction, CanUD showed significant enrichment in six IT cortical layer cell types, and PAU showed significant enrichment in seven cell types, including IT cortical neurons and MSN subtypes (**Table S5**). No associations survived Bonferroni correction for any phenotype (p < 1.02 × 10 ³ for 49 cell types tested).

**Figure 3:**
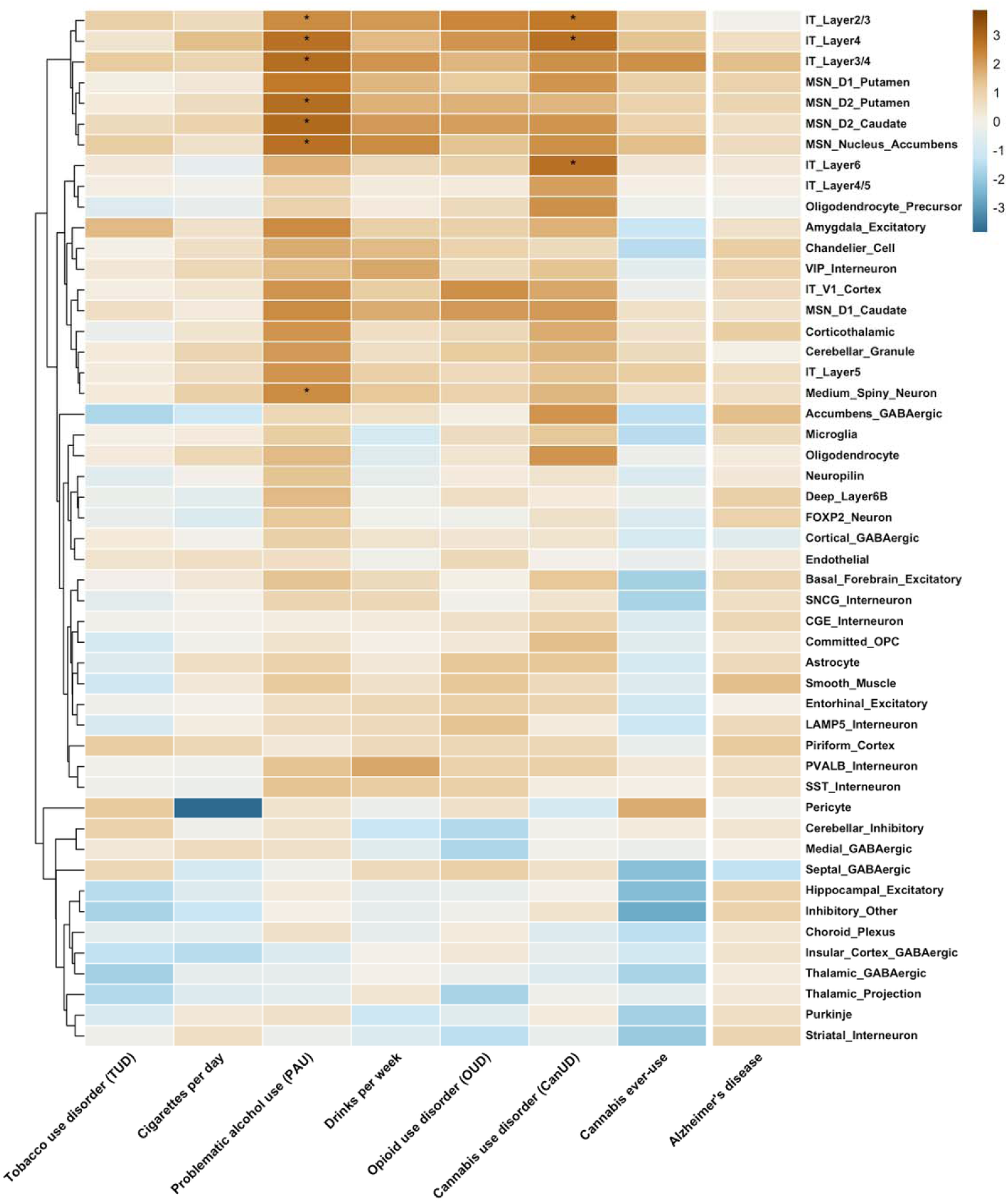
LDSC enrichment Z-scores across consolidated snATAC-seq cell-type categories for seven substance use phenotypes identified brain region cell-type specific enrichment patterns for Cannabis Use Disorder and Problematic Alcohol Use. Brown indicates positive enrichment; blue indicates negative enrichment. Rows are hierarchically clustered. Original Li snATAC-seq annotations were grouped into broader cell-type categories for visualization, with heatmap colors representing the mean enrichment Z-score across the original annotations within each category. * indicates that at least one original annotation within the corresponding consolidated category was significant after FDR correction (FDR < 0.05). No associations survived Bonferroni correction across the 109 original snATAC-seq annotations (Bonferroni *p* < 4.59 × 10⁻□). Among the seven phenotypes, CanUD and PAU exhibited the strongest cell-type-specific enrichment, primarily across cortical intratelencephalic neurons and striatal medium spiny neuron subtypes, whereas TUD showed no significant enrichment in the snATAC-seq analysis. In contrast, Alzheimer’s disease, included as a negative control, showed a distinct enrichment pattern.

At the nominal significance threshold (p < 0.05), the most widespread enrichment was observed for PAU, which showed positive enrichment in 45 cell types, predominantly cortical excitatory IT neuron subtypes across multiple layers (ITL2/3, ITL4, ITL5, and ITL6) as well as striatal subpopulations, including MSN_1, MSN_2, and D2CaB. Drinks per week showed enrichment in 27 cell types, again concentrated in cortical IT neurons and striatal subtypes. CanUD showed nominal enrichment in 35 cell types, with the strongest signals in upper and deep layer cortical IT neurons. In contrast, TUD showed no nominally significant positive enrichment in any cell type, and cannabis ever use and cigarettes per day showed only three nominally significant associations each, suggesting limited power or a weaker signal in these phenotypes in the ATAC-seq annotation system. Across phenotypes showing any nominal enrichment, the direction of effect was consistently positive in excitatory neuronal populations, mirroring the pattern observed in the snRNAseq analysis.

### *Correlations between snRNAseq and snATACseq enrichment and between substance* use and use disorders

To determine whether enrichment signals were consistent across the two annotation modalities, we computed Pearson correlations between matched snRNAseq and snATACseq enrichment Z-scores across 44 cell-type pairs for each phenotype (**Figure 4A**, **Table S6**). Drinks per week showed the strongest correlation (r = 0.56, p = 7.67 × 10⁻), followed by PAU (r = 0.53, p = 1.90 × 10⁻) and CanUD (r = 0.41, p = 0.006), indicating that cell types showing greater heritability enrichment in the snRNAseq data also tended to show greater enrichment in the snATACseq data for these phenotypes. OUD showed a modest positive correlation that did not reach significance (r = 0.28, p = 0.061), and cigarettes per day and cannabis use showed weak non-significant correlations (r = 0.20 and r = 0.18, respectively). Notably, TUD showed no evidence of concordance between annotation systems (r = −0.04, p = 0.79), consistent with its distinct enrichment profiles observed in the snRNAseq (Figure **2**) and its lack of significant enrichment in snATACseq analyses (**Figure 3**).

**Figure 4:**
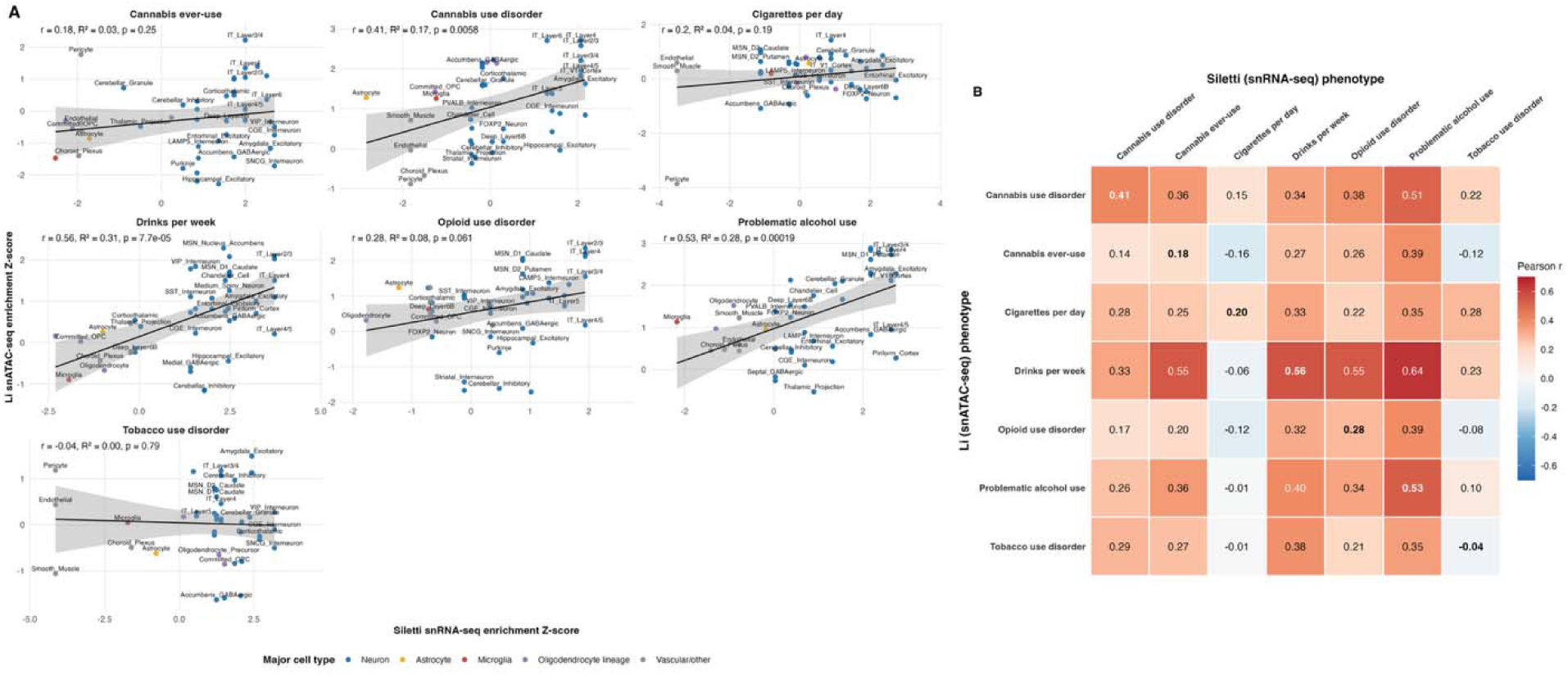
(A) Scatter plots of LDSC enrichment Z-scores for the two annotation systems for each SUD phenotype. Each point represents a matched cell-type annotation between the Siletti snRNA-seq and Li snATAC-seq datasets, with points colored by major cell-type category. The black line represents the linear regression fit with a 95% confidence interval. Pearson correlation coefficients (*r*), corresponding R² values, and *p*-values are shown in each panel. Cross-modality concordance was strongest for drinks per week, problematic alcohol use, and cannabis use disorder, whereas tobacco use disorder showed little evidence of cross-modality concordance. (B) Pearson correlations across SUDs and data modalities (snATACseq and snRNAseq). Within-phenotype, cross-modality correlations are presented along the diagonal. Bold diagonal values represent matched-phenotype cross-modality correlations, whereas off-diagonal values represent mismatched-phenotype cross-modality correlations. Red indicates positive correlations, and blue indicates negative correlations.

We also examined correlations across modalities and substance use phenotypes (**Figure 4B**); these cross-phenotype, cross-modality correlations tended to be of similar magnitude to the within-phenotype, cross-modality correlations described above. One exception was cigarettes per day, which generally showed weaker and even negative cross-modality correlations with other phenotypes. Interestingly, drinks per week snATACseq enrichments were equally or more strongly correlated with cannabis use (r = 0.55), OUD (r = 0.55), and PAU (r = 0.64) snRNAseq enrichments than with drinks per week’s own snRNAseq enrichments (r = 0.56). Because of the relatively strong cross-phenotype correlations, we fit an empirical background distribution of cross-phenotype, cross-modality correlations. The off-diagonal (mismatched-phenotype) Pearson correlations from the 7×7 cross-phenotype correlation matrix (**Figure 4B**) were Fisher z-transformed and fit to a normal distribution, which showed no significant departure from normality (Shapiro-Wilk W = 0.955, p = 0.10). The 95th and 99th percentiles of the fitted background distribution corresponded to Pearson correlations of r = 0.538 and r = 0.629, respectively (**Figure S1**). Among the seven matched within-phenotype correlations, only drinks per week (r = 0.56) exceeded the 95th-percentile threshold, with PAU (r = 0.53) falling just below this threshold. None exceeded the 99th-percentile threshold.

To further explore concordance between substance use and substance use disorder phenotypes, we examined within-modality correlations between disorder and corresponding use phenotypes (e.g., TUD vs. cigarettes per day, PAU vs. drinks per week, and CanUD vs. cannabis ever-use; OUD was not tested due to the absence of a corresponding use phenotype) (**Table S7**). These within-modality correlations were generally stronger than the cross-modality correlations described above, particularly in the snRNA-seq annotation (PAU-drinks per week: r = 0.91, p = 1.7 × 10 ¹²; CanUD–cannabis ever-use: r = 0.75, p = 1.5 × 10; TUD-cigarettes per day: r = 0.70, p = 1.1 × 10), with a similar but attenuated pattern observed in the snATAC-seq annotation (PAU–drinks per week: r = 0.61; CanUD–cannabis ever-use: r = 0.56; TUD-cigarettes per day: r = 0.37; all p < 10). These findings indicate that substance use behaviors and their corresponding use disorders show broadly similar cell-type enrichment patterns across both annotation modalities, although concordance was consistently strongest for alcohol-related phenotypes and weakest for tobacco-related phenotypes.

## Discussion

In this study, we leveraged cell-type-specific heritability enrichment analysis to connect the genetic architecture of substance use behaviors and SUDs to specific brain cell types, using two independent single-cell datasets profiling gene expression and chromatin accessibility across the adult human brain. Our results reveal a consistent pattern of enrichment concentrated in excitatory neuronal populations, most prominently upper-layer intra-telencephalic, amygdala-excitatory, hippocampal, and striatal neurons across multiple phenotypes and for both gene expression and chromatin accessibility (**Figure 5)**, although enrichments surviving FDR correction were confined to drinks per week and TUD in the snRNAseq analysis and CanUD and PAU in the snATACseq, and few associations survived the more stringent Bonferroni correction. These findings extend prior work implicating neuronal populations in SUD heritability^1^ and provide higher cellular resolution by leveraging human single-nucleus data rather than mouse transcriptome atlases or bulk tissue.

**Figure 5:**
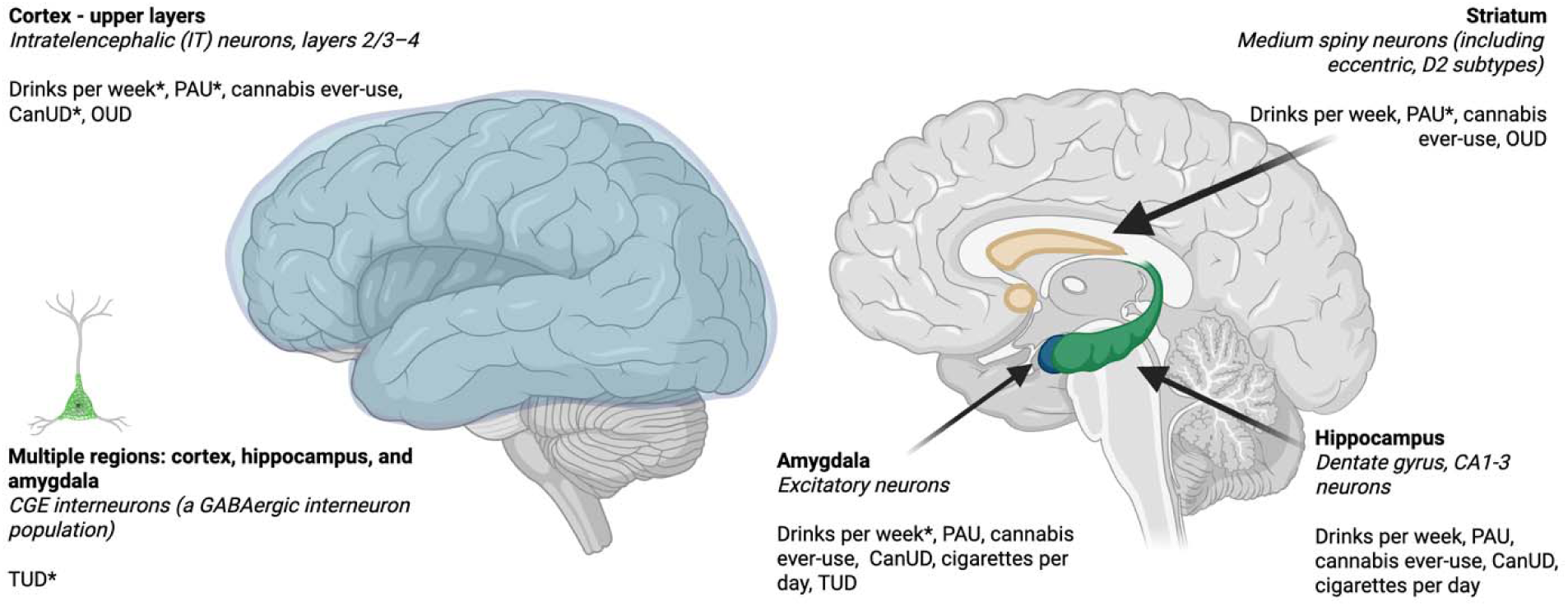
Brain regions implicated in cell-type specific heritability enrichment for substance use and substance use disorders. Brain schematics illustrating the anatomical locations of neuronal populations identified in cell-type specific heritability enrichment analyses using annotations developed from snRNA-seq and snATAC-seq data. Left: Lateral view highlighting cortical regions containing upper layer intratelencephalic (IT) neurons and CGE interneurons, a GABAergic interneuron population distributed across cortex, hippocampus, and amygdala. Right: Sagittal view highlighting the amygdala (excitatory neurons), striatum (medium spiny neurons, including D2 and eccentric subtypes), and hippocampus (dentate gyrus and CA1-3). *Indicates phenotypes that reached FDR-corrected significance (FDR < 0.05); all other listed phenotypes were nominally significant (p < 0.05, uncorrected).

The convergent enrichment of SUD heritability in upper layer intratelencephalic and amygdala excitatory neurons across multiple phenotypes is consistent with the established neurobiology of addiction^18,19^. Cortical intratelencephalic neurons^20^, which project extensively within the cortex and to the striatum, are well positioned to contribute to the cognitive and motivational processes dysregulated in addiction, including impaired impulse control and compulsive drug seeking^21^.

The consistent enrichment (at p < 0.05) of amygdala excitatory neurons across six of seven phenotypes is particularly noteworthy given the well-characterised role of the basolateral amygdala in encoding drug-associated memories and in motivating reward-seeking behaviour through its glutamatergic projections to the nucleus accumbens^22^. The recurrent enrichment of medium spiny neurons, both eccentric and canonical, across alcohol, cannabis, and opioid phenotypes further aligns with the central role of striatal dopaminergic circuitry in processing reward and reinforcing drug-taking behaviour. These findings are consistent with a recent study integrating the same human brain atlas from Siletti et al. with GWAS summary statistics across psychiatric disorders and substance use-related traits, which similarly identified enrichment of excitatory cortical neurons, medium spiny neurons, and cortical interneurons across several phenotypes, while amygdala excitatory neurons showed more selective enrichment^23^. However, that study examined only three substance-related phenotypes (problematic alcohol use, alcohol consumption, and smoking initiation) within a broader analysis of psychiatric and brain phenotypes, whereas the present study focuses specifically on substance use and SUDs. The overlap between these findings and our results provides independent support for the involvement of cortico-striatal-limbic neuronal populations in the genetic architecture of substance use disorders.

Notably, several of the cell types most consistently enriched in our analyses are also associated with genes with well-established roles in addiction neurobiology. Medium spiny neurons, particularly the D2 (indirect pathway) subtype, are characterized by expression of *DRD2*, which encodes the dopamine D2 receptor, a key component of striatal dopamine signaling implicated in addiction-related reward, reinforcement, and habit circuitry^19,24^. This convergence is notable given that *DRD2* has also been implicated in multivariate GWAS of shared genetic liability across SUDs^2^. Similarly, *OPRM1*, which encodes the μ-opioid receptor and is a key pharmacological target in OUD treatment, was implicated by genome-wide significant variants in the OUD GWAS^10^, providing further convergence between our cell-type enrichment findings and established genetic and pharmacological evidence for SUDs.

Interestingly, the predominance of excitatory neuronal enrichments observed in our analyses contrasts with postmortem transcriptomic studies of SUDs, which frequently report prominent alterations in microglial, astrocytic, and other neuroimmune pathways^25,26^. Another study found that polygenic liability for alcohol use disorder (AUD) moderated the response of microglia to ethanol exposure, suggesting some interplay between heritable risk for AUD, inflammation, and response to alcohol^27^. The apparent discrepancy in our results may reflect distinct biological processes captured by these complementary approaches, as well as the nonspecificity of neuroimmune responses. Cell-type-specific heritability enrichment identifies neuronal populations through which inherited genetic risk is preferentially mediated, whereas postmortem transcriptomic studies primarily capture molecular adaptations arising from chronic substance exposure. Together, these findings support a model in which genetic susceptibility to SUDs is mediated predominantly through neuronal circuits involved in reward, learning, and executive functioning, while prolonged substance exposure subsequently elicits glial activations and neuroimmune responses that contribute to disease progression^19,25^. In contrast, Alzheimer’s disease exhibited a markedly different enrichment profile, with heritability predominantly concentrated in microglia rather than excitatory neuronal populations, supporting the specificity of the neuronal enrichment observed for SUDs.

TUD showed a strikingly divergent pattern relative to the other SUDs and substance use phenotypes examined. In general, disorder phenotypes and their corresponding consumption phenotypes generally showed broadly similar enrichment profiles-for example, PAU and drinks per week both showed strong consistent excitatory neuron enrichment, as did CanUD and cannabis ever-use. TUD and cigarettes per day were notable exceptions, showing more limited and less consistent enrichment signals compared to the alcohol and cannabis phenotypes. In the snRNAseq annotation, cigarettes per day showed nominal enrichment concentrated in amygdala excitatory and hippocampal CA1-3 neurons, with more modest signals in deep-layer corticothalamic/6b and upper-layer intratelencephalic neurons **(Figure 1)**, broadly overlapping with the excitatory populations enriched for other consumption phenotypes rather than mirroring TUD’s inhibitory-shifted profile. This could be due, at least in part, to greater specificity of the molecular target of nicotine as opposed to ethanol; it could be that TUD enrichments reflect highly specific neuronal nicotinic response pathways, while alcohol-related enrichments reflect ethanol’s broader metabolic effects on nearly all populations of neurons^28^. In the snATACseq annotation, cigarettes per day showed only weak, largely non-significant enrichment across cell types **(Figure 3)**. In contrast to the predominantly excitatory neuronal enrichment observed across other SUDs, TUD showed a pattern shifted towards inhibitory neuronal populations, with its strongest enrichment signal in CGE interneurons, a population of GABAergic inhibitory neurons. Among all phenotypes examined, TUD showed the weakest concordance between snRNAseq and snATACseq enrichment profiles (r = −0.04), in contrast to the significant positive correlations observed for drinks per week (r = 0.56), PAU (r = 0.53), and CanUD (r = 0.41).

Consistent with this distinction, the cell-type enrichment profiles of TUD and cigarettes per day were positively correlated but less strongly than the corresponding alcohol and cannabis phenotype pairs (snRNA-seq: *r* = 0.70 and snATAC-seq: *r* = 0.37, compared to *r* = 0.91 and *r* = 0.61 for alcohol phenotypes). Interestingly, TUD and cigarettes per day showed markedly different cell-type enrichment patterns despite both capturing tobacco-related phenotypes and being modestly genetically correlated at the genome-wide level (rg = 0.44, SE = 0.03)^1^, suggesting that the genetic architecture of clinically defined tobacco use disorder may differ from that of tobacco consumption behaviour in the specific neuronal populations through which genetic risk is mediated.

Several limitations should be considered when interpreting these findings. First, all GWAS included in this study were conducted primarily in individuals of European ancestry, limiting the generalizability of our findings to other ancestral populations. Second, the statistical power of the included GWAS varied across substance use and SUD phenotypes. In particular, the OUD GWAS, which had the smallest effective sample size among the disorders examined, did not yield significant cell-type enrichments, suggesting that limited statistical power may have reduced our ability to detect true biological signals. As larger and more ancestrally diverse GWAS become available, particularly for OUD, they may provide greater power to detect and validate cell-type enrichments associated with SUD risk. Third, our analyses did not incorporate epigenetic data, such as CpG methylation, nor did they consider region-specific transcriptomic or epigenomic profiles, which may provide additional insight into the molecular mechanisms underlying addiction susceptibility. Finally, although our findings support a predominantly neuronal genetic architecture for SUDs, integrating genetic enrichment analyses with postmortem transcriptomic and other multi-omic datasets will be important for distinguishing genetic liability from molecular changes that occur as a consequence of chronic substance use.

Our findings demonstrate that embedded within the common variant genetic architecture underlying substance use and SUD liability are predispositional differences in neuronal gene expression and chromatin accessibility. While some of the enriched cell types that we identified were also implicated in postmortem brain expression and multiome assays, the limited extent of microglial enrichment in our data and the corresponding weak enrichment of GWAS signal in published postmortem data from SUD decedents suggest a potential mechanistic divergence. Taken together, these findings implicate pre-existing differences in neuronal gene expression in brain regions fundamental to the development of addictions with convincing support that GWAS effectively captures this variability. The role of glial mechanisms that predominate in postmortem gene expression studies that compare SUD and control samples is consistent with the potential neuroinflammatory consequence of prolonged exposure to psychoactive substances. This neuronal and glial interplay provides a preliminary framework for the development of experimental studies aimed at dissecting the precise mechanism of drug action (e.g., induction of pluripotent stem cells into neurons vs. glia to study differing aspects of addiction development).

## Data Availability

The datasets analyzed in this study are available from publicly accessible repositories and resources as described in the Methods. GWAS summary statistics for the substance use and substance use disorder phenotypes were obtained from the original studies and their respective repositories. Single-nucleus RNA-sequencing and chromatin accessibility data were obtained from the previously published datasets described in the Methods.

## Author Contributions

AA, ECJ, and NS contributed to the conception of the study. NS and ECJ contributed to data analysis. YEL and ZP advised on study design and statistical analyses. AA, ECJ, NS, YEL, and ZP contributed to the interpretation of the results. NS and ECJ drafted the manuscript, and AA provided critical edits. All authors reviewed the manuscript draft, provided feedback, and gave approval of the final manuscript.

## Acknowledgements

Generative artificial intelligence (Claude, Anthropic) was used during manuscript preparation to assist with language editing and improve clarity and readability. All scientific content, interpretations, and final manuscript text were reviewed and verified by the authors. This work utilized the High Throughput Computing Facility at the Center for Genome Sciences and Systems Biology at Washington University in St. Louis.

## Supplementary Information

Supplementary information is available at MP’s website.

## Supplementary Information

**Supplementary Figure 1.**
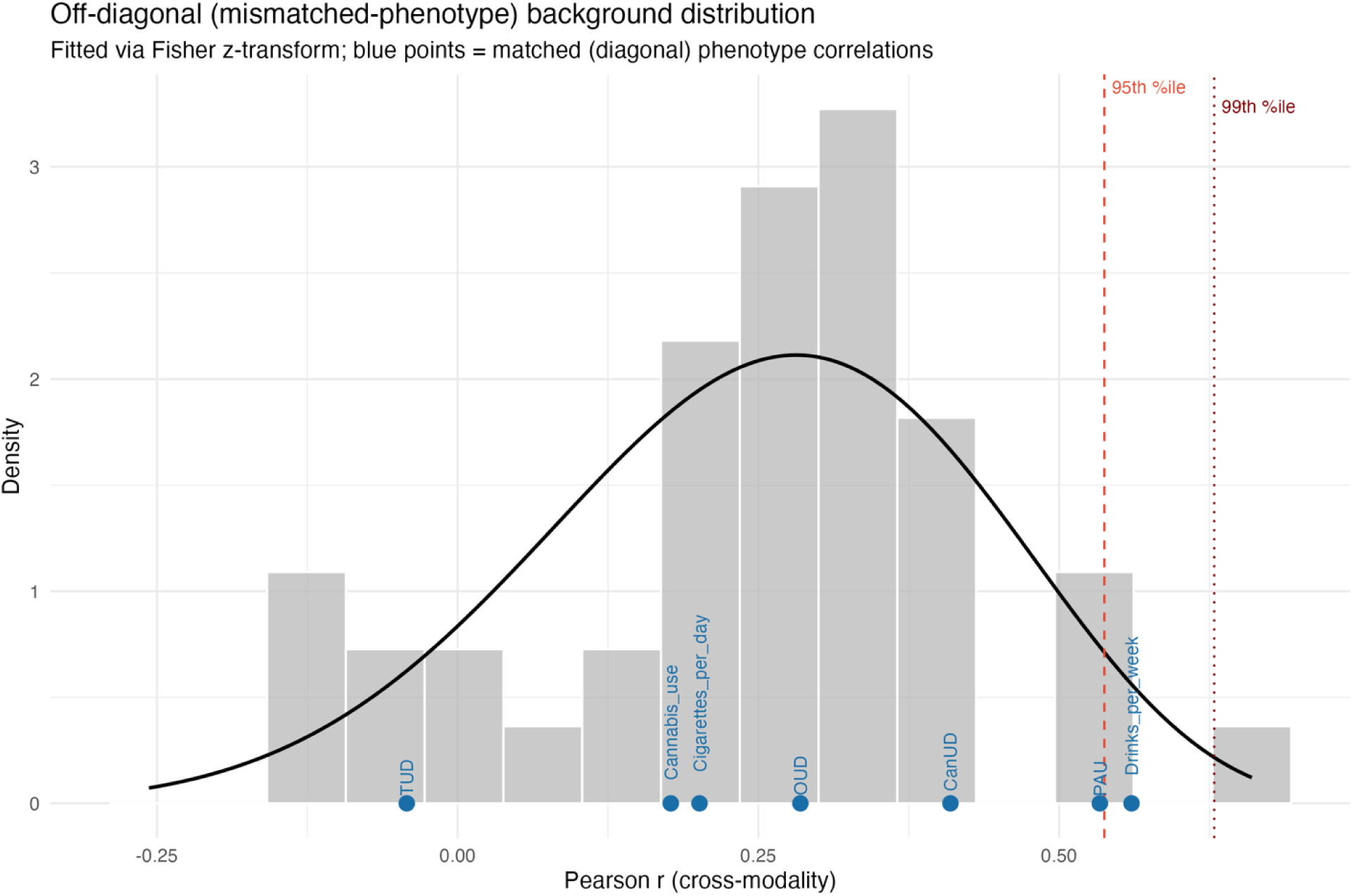
Empirical background distribution of cross-phenotype, cross-modality correlations. Off-diagonal (mismatched-phenotype) Pearson correlations from the 7×7 cross-phenotype correlation matrix (Figure 4B) were Fisher z-transformed and fit to a normal distribution (Shapiro-Wilk W=0.955, p=0.10, supporting normality). The solid black line shows the fitted density; dashed and dotted vertical lines indicate the 95th- and 99th-percentile cutoffs (r=0.538 and r=0.629, respectively), back-transformed to the Pearson correlation scale. Blue points show the observed within-phenotype (matched) cross-modality correlation for each of the seven substance use/SUD phenotypes. Only drinks per week exceeded the 95th-percentile threshold.

