## Supplementary Figure for "Cell type specific enrichment of substance use and substance use disorder heritability"

**Supplementary Figures for Cell type specific enrichment of substance use and substance use disorder heritability**

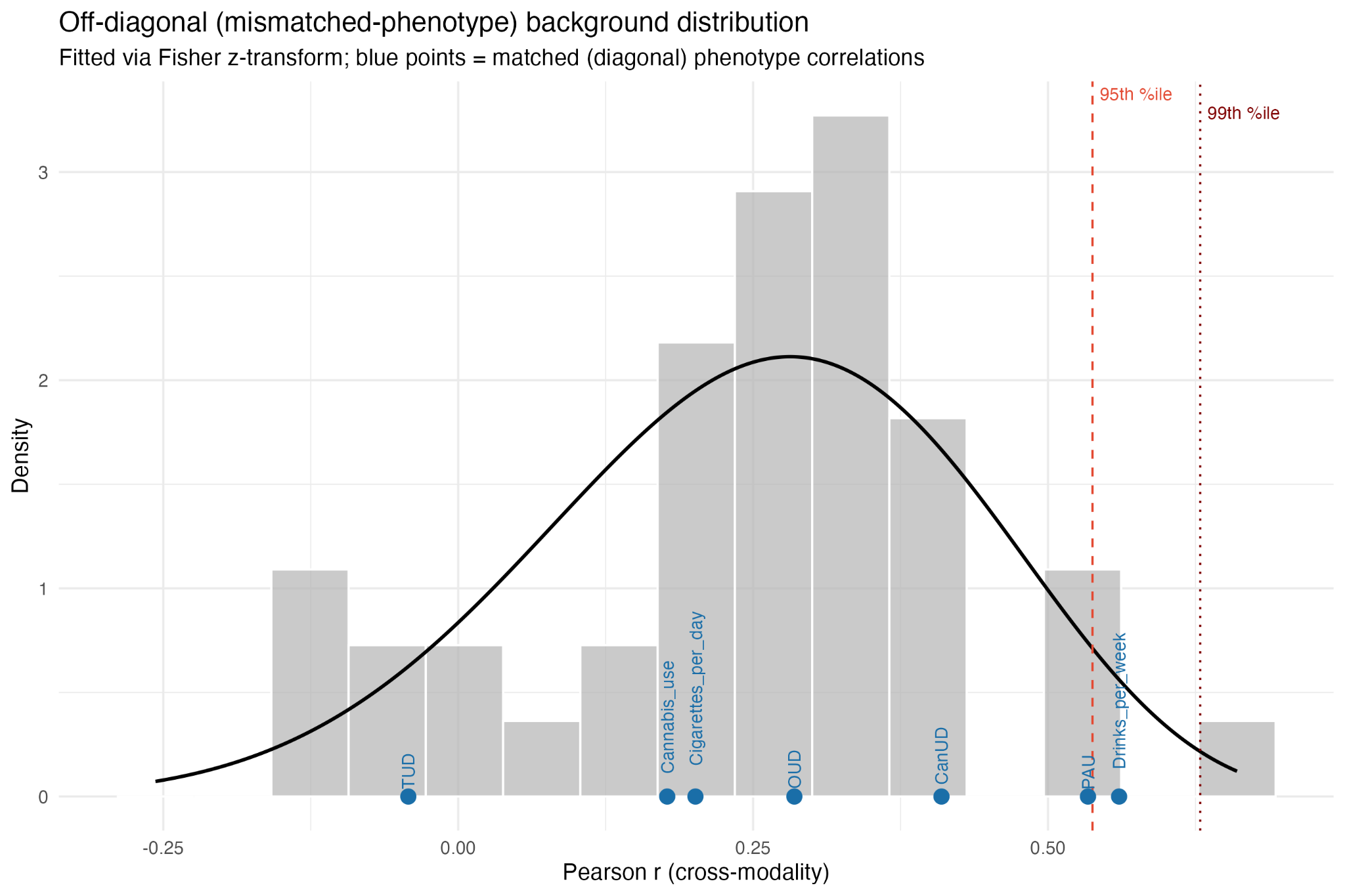

**Supplementary Figure 1.** Empirical background distribution of cross-phenotype, cross-modality correlations. Off-diagonal (mismatched-phenotype) Pearson correlations from the 7×7 cross-phenotype correlation matrix (**Figure 4B**) were Fisher z-transformed and fit to a normal distribution (Shapiro-Wilk W=0.955, p=0.10, supporting normality). The solid black line shows the fitted density; dashed and dotted vertical lines indicate the 95th- and 99th-percentile cutoffs (r=0.538 and r=0.629, respectively), back-transformed to the Pearson correlation scale. Blue points show the observed within-phenotype (matched) cross-modality correlation for each of the seven substance use/SUD phenotypes. Only drinks per week exceeded the 95th-percentile threshold.
